# Prenatal phenotype of *PNKP*-related primary microcephaly associated with variants in the FHA and Phosphatase domain

**DOI:** 10.1101/2021.09.25.21261035

**Authors:** Sonja Neuser, Ilona Krey, Annemarie Schwan, Rami Abou Jamra, Tobias Bartolomaeus, Jan Döring, Steffen Syrbe, Margit Plassmann, Stefan Rohde, Christian Roth, Helga Rehder, Maximilian Radtke, Diana Le Duc, Susanna Schubert, Luis Bermúdez-Guzmán, Alejandro Leal, Katharina Schoner, Bernt Popp

## Abstract

Biallelic *PNKP* variants cause heterogeneous disorders ranging from neurodevelopmental disorder with microcephaly/seizures to adult-onset Charcot-Marie-Tooth disease. To date, only postnatal descriptions exist.

We present the first prenatal diagnosis of *PNKP*-related primary microcephaly. Detailed pathological examination of a male fetus revealed micrencephaly with extracerebral malformations and thus presumed syndromic microcephaly. A recessive disorder was suspected because of previous pregnancy termination for similar abnormalities in a sibling fetus. Prenatal trio exome sequencing identified compound-heterozygosity for the *PNKP* variants c.498G>A, p.[(=),0?] and c.302C>T, p.(Pro101Leu). Segregation confirmed both variants in the sibling fetus. Through RNA analyses, we characterized skipping of exon 4 affecting the *PNKP* Forkhead-associated (FHA) and Phosphatase domains (p.Leu67_Lys166del) as the predominant effect of the c.498G>A variant. We retrospectively investigated two unrelated individuals diagnosed with biallelic *PNKP*-variants to compare prenatal/postnatal phenotypes. Both carry the same splice-donor variant c.1029+2T>C *in trans* with a variant in the FHA domain (c.311T>C, p.(Leu104Pro) and c.151G>C, p.(Val51Leu), respectively). RNA-seq showed complex splicing events for c.1029+2T>C and c.151G>C. Computational modelling and structural analysis revealed significant clustering of missense variants in the FHA domain, with some variants potentially generating structural damage.

Our detailed clinical description extends the *PNKP*-continuum to the prenatal stage. Investigating possible *PNKP*-variant effects using RNA and structural modelling, we highlight the mutational complexity and exemplify a framework for variant characterization in this multi-domain protein.

## INTRODUCTION

Microcephaly, or rather micrencephaly (abnormally small brain) in the narrow sense, is defined as an occipital frontal circumference (OFC) below -2 SD of the mean for (gestational) age and sex and can occur in isolated form or in a syndromic context (1). If detected prenatally, it is classified as primary microcephaly (PM) in contrast to secondary microcephaly (SM) developing after birth. Infections, traumata, ischemic events, exposure to teratogens and genetic disorders are possible etiologies (1,2). As head growth - among many other factors - depends on normal neuronal tissue proliferation, requiring continuous cell division, several genetic neurodevelopmental and neurodegenerative disorders in this context are caused by variants affecting DNA repair genes, highlighting the importance of the pathways in neurogenesis (3).

The Polynucleotide kinase 3’-phosphatase (PNKP) has a dual Kinase/Phosphatase function and is involved in the repair of both single- and double-strand DNA breaks (4,5). In 2010, biallelic pathogenic *PNKP* variants were reported to cause “microcephaly, seizures and developmental delay” (MCSZ; MIM# 613402) (6). Over time, three additional neurological and neurodevelopmental syndromes have been associated with disease-causing variants in *PNKP*. These range from “childhood-onset ataxia with oculomotor apraxia type 4” (AOA4; MIM# 616267) (7) to “developmental and epileptic encephalopathy” (DEE10; MIM# 613402) (8) and “adult-onset Charcot-Marie-Tooth disease, type 2B2” (CMT2B2; MIM# 605589) (9,10). To date, exclusively postnatal observations are covered in literature.

Most pathogenic *PNKP* variants described so far are either truncating or located in the C-terminal Kinase domain (11). While genotype-phenotype correlations have been attempted and C-terminal variants have been implied to cause the milder adult-onset diseases, no clear relation could yet be established. Instead, it has even been postulated that the pathogenic variants observed present with rather mild mutational effects, due to survivorship bias, and more damaging variants would result in non-viability (11).

Here, we describe the first prenatal identification of biallelic *PNKP* variants affecting the region between the N-terminal Forkhead-associated (FHA) domain and the Phosphatase domain causative for severe early onset of PM. We provide detailed descriptions based on prenatal imaging and syndrome-oriented fetal autopsies of two affected sibling fetuses, we compare the fetal phenotype with two individuals with *PNKP*-associated disorder diagnosed previously and give a review of similar cases in the literature. Additionally, we performed extensive RNA analyses to characterize aberrant splicing of identified variants and used structural modelling to investigate the effects of missense variants.

## MATERIALS AND METHODS

### Ethics approval

The study adheres to the principles set out in the Declaration of Helsinki. The Ethical Committee of the Medical Faculty, Leipzig University approved genetic testing in a research setting for all probands within the study. Written consent of the parents to publish genetic and clinical data, as well as prenatal images and postmortem photographs (P1, P2), sonography (P1, P2), and magnetic resonance imaging (MRI) images (P1, P3 and P4) was received and archived by the authors.

### Genetic analyses and review of *PNKP* variants

P2 and both his parents underwent trio-exome sequencing. In P3 and P4, clinical exome sequencing (CES) was performed. Segregation was confirmed through Sanger sequencing in all. Technical details and primer sequences are provided in Supplementary notes. All *PNKP* variants have been submitted to ClinVar (File S3 (12) sheet “PNKP_variants”).

A PubMed search using the term “PNKP” identified 38 distinct *PNKP* variants from 21 publications (searched on 2020-12-15, detailed literature references in File S3 (12) sheet “PNKP_variants”). Variants were standardized to the *PNKP* reference transcript NM_007254.3 (GRCh37/hg19) using Mutalyzer 2.0.32 and annotated as described previously (13) (see File S3 (12) sheet “PNKP_variants”). All *PNKP* variants described here were classified following current American College of Medical Genetics and Genomics (ACMG) guidelines (14).

### Clinical data collection

We used a questionnaire for retrospective phenotype analysis in which clinical terms were standardized using the Human Phenotype Ontology (HPO) terminology (15) based on a review of previously reported clinical associations in *PNKP*-disorders (6–10). The sheet was sent for evaluation to the pediatric neurologist or pathologist, respectively and data from available clinical reports were added. Pre- and postnatal anthropometric measurements were compared to standard values according to Potter et Craig (16), or WHO child growth standards (17). Comprehensive results are shown in File S2 (12) sheet “clinical” and Figure S1.

### Fetal autopsy and RNA extraction from fetal tissue

Fetal pathological examination of P1 and P2 was performed as previously described (18), detailed information is shown in File S1. Cryopreserved native skeletal muscle tissue of P2 was processed with QIAshredder (Qiagen, Hilden, Germany) and RNA was extracted according to the manufacturer’s protocol (RNeasy Mini, Qiagen, Hilden, Germany).

### RNA analyses

In family 1 we performed RT-PCR as described previously (19) using PAXgene RNA (Becton Dickinson, Franklin Lakes, NJ) in the parents and fetal skeletal muscle RNA derived cDNA. In family 3, we performed RNA-seq from PAXgene RNA using the TruSeq RNA Library Prep Kit v2 according to the manufacturer’s instructions and paired-end sequencing on an Illumina NextSeq platform. Bioinformatic workup included an established pipeline from our institute. In brief, reads were demultiplexed, adapters trimmed, overrepresented sequences removed before we aligned the remaining reads to the hg38 reference using STAR aligner (20). We visualized and inspected the alignments for aberrant splicing as described previously (21) and applied iREAD (22) to quantify the visually observed intron retention events. Plotting of Figure 3 was performed with R using the packages “ggplot2”, “Gviz” and “trackViewer” and Inkscape was used to adjust figure components. Detailed procedures including primer sequences, reagents and software versions are provided in the Supplementary notes.

### Analysis of missense variant spectrum

Analysis of disease-associated missense variants in the linear protein representation, clustering analysis in 3D and structural modelling of missense variants using the crystal structure 2BRF (23) was performed as described previously (11,13,19) and is detailed in the Supplementary notes.

## RESULTS

### Prenatal phenotype in two sibling fetuses

In the pregnancy of a healthy non-consanguineous couple, routine sonography revealed microcephaly, abnormal skull shape and microretrognathia in the male fetus (P2). Follow-up ultrasound examinations displayed progression of the described anomalies. Amniocentesis for genetic testing (trio ES) was performed. Severe fetal anomalies and supposing genetic background caused the parents to decide for termination of pregnancy.

A previous pregnancy had been terminated after prenatal imaging had confirmed multiple anomalies in a female fetus (P1). Ultrasound had shown microcephaly, asymmetric skull shape, abnormal brain development, cerebellar hypoplasia, cataract of both eyes, and facial abnormalities. Prenatal MRI confirmed microcephaly, large supratentorial defects of brain parenchyma in occipital, parietal and frontal regions, severe cerebellar hypoplasia, dilatation and fusion of both lateral ventricles (Figure 1C). Corpus callosum and septum pellucidum were not determinable. Brainstem and thalamus appeared normal, basal ganglia were not accessible. Bulbi of the eyes differed in size and signal. Other organs were described as normal.

**Figure 1.**
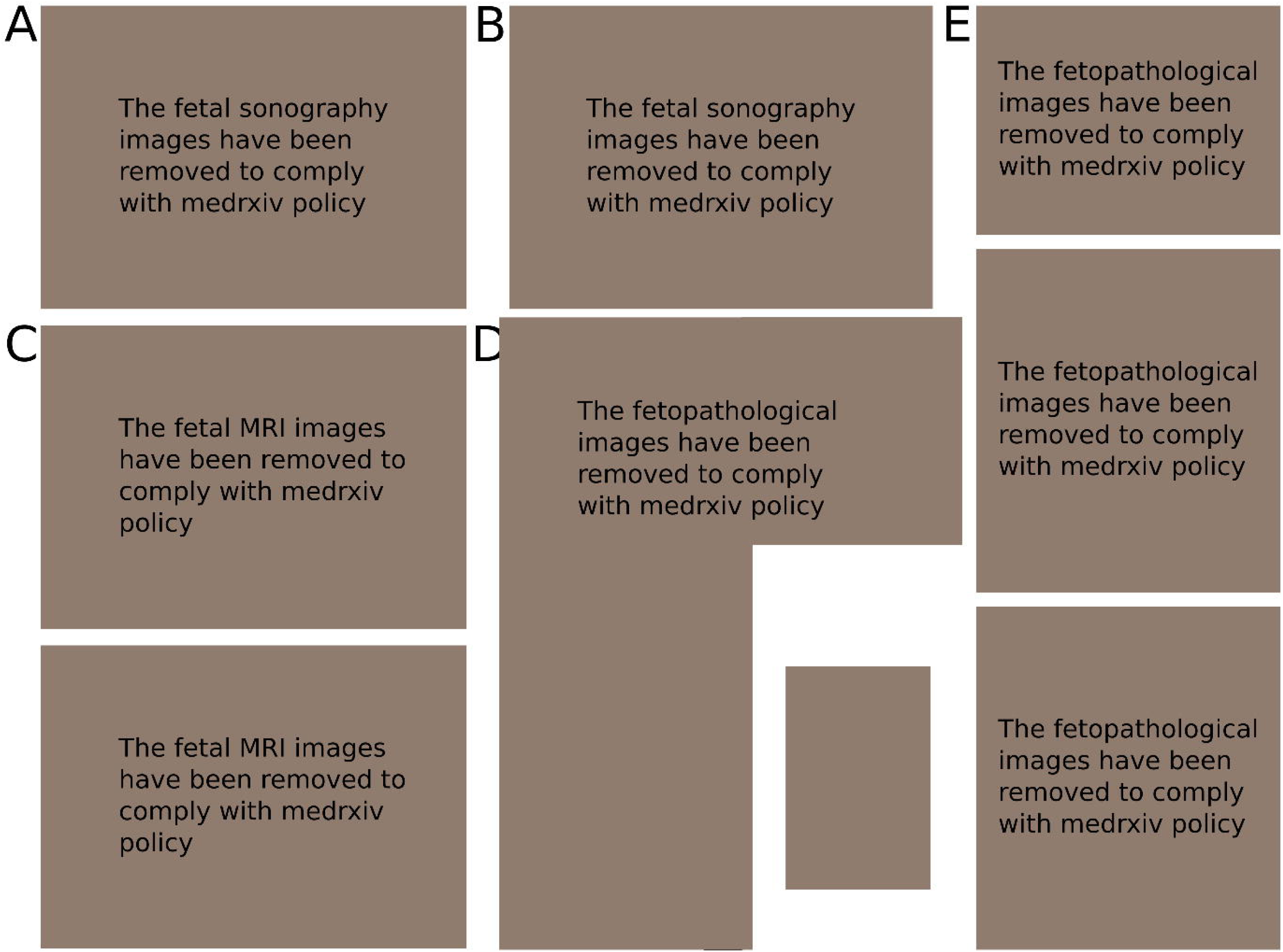
Prenatal Clinical and Autopsy Results. **(A)** and **(B)** fetal sonography images removed to comply with medrxiv policy **(C)** fetal MRI images removed to comply with medrxiv policy **(D)** and **(E)** Postmortem examination images removed to comply with medrxiv policy

Both fetuses were examined after TOP. The female fetus P1 measured 24.0 cm crown to heel length (standard value: 26.2±3.6 cm (16) having a weight of 301 g (standard value: 353±125 g (16)). OFC was 15.0 cm (−4.3 SD; 5^th^ percentile at GA: 17.3 cm (24)). Autopsy presumed severe cerebral parenchymatous defects, profound hypoplasia of posterior cranial fossa and confluence of the side ventricles. The findings were initially interpreted as arhinencephaly/holoprosencephaly. Further, extracerebral anomalies were not assessable, also because of autolysis. Placenta appeared hypotrophic with signs of insufficiency.

The sibling fetus P2 was almost age-appropriate in terms of crown to heel length of 18.8 cm and weight (18.8 cm resp. 124.3 g) (standard values: 20 cm and 150 g), but had extremely small OFC (11.3 cm, -5.85 SD; 5^th^ percentile at this GA: 14.1 cm (24). Fetal autopsy confirmed severe micro- and brachycephaly, short receding forehead, narrow fontanelles and associated facial dysmorphisms including hypertelorism, anteverted nares, long philtrum, small upper lip and small outer ears. Moreover, the fetus showed contractures according to early manifestation of arthrogryposis. Brain volume was reduced (about 40% of standard value, GA norm (16)) with slight enlargement of the ventricles. The frontal lobes were hypoplastic, occipital lobes were shortened and appeared wing-like. Temporo-parietal lobulation and corpus callosum were missing. Cerebellum was hypoplastic with a diameter of 1.2 cm (5^th^ percentile for GA: 1.6 cm (24)). All examined brain sections appeared histologically normal. The findings were interpreted as micrencephaly without neuronal migration disorder or structural malformations. Examination of the eyes revealed discrete anisophthalmia and anisocoria with partial lens luxation due to dysplasia of the iris and persistent hyaloid artery of the left eye. There were no signs for infectious, toxic or hypoxic influences and no further organ abnormalities. Fetal autopsy results of P2 were suspected as monogenic syndromic type of microcephaly.

### Genetic analyses

Initial genetic investigations in P1, including conventional karyotyping, chromosomal microarray analysis and a targeted holoprosencephaly sequencing panel (15 genes: *CDON, DHCR7, DLL1, EYA4, FBXW411, FGF8, GAS1, GLI2, GLI3, PTCH1, SHH, SIX3, SMAD2, TGIF1, ZIC2*), were unremarkable. The recurrent pattern of PM in two successive pregnancies suggested a recessive syndromal type of PM. Thus, trio-ES was initiated after amniocentesis of P2. This analysis revealed the compound heterozygous *PNKP* variants c.498G>A, p.[(=),?] and c.302C>T, p.(Pro101Leu). The variant c.498G>A is formally annotated as synonymous (p.(=)), but affects the last base of exon 4 and is predicted to disrupt the splice donor motif (p.(?)) with simple skipping of exon 4 resulting in an in-frame deletion between the FHA and the Phosphatase domains. The variant c.302C>T causes a proline to leucine missense change at the amino acid (AA) position 101 in the FHA domain. Both were initially classified as variants of unknown significance (VUS) according to ACMG recommendations (criteria: PM2, PP3). Subsequent segregation analysis in an archived amniotic fluid sample confirmed these two variants in compound-heterozygous state in the first affected fetus P1. Despite this cosegregation evidence supporting pathogenicity the variants’ classification remained VUS.

### Postnatal phenotype in two unrelated individuals

*This case report has been removed from the preprint version to comply with medrxiv policy. Please see the published version or contact the authors if you are interested in this information*.

CES identified the two heterozygous *PNKP* variants c.1029+2T>C, p.(?) and c.311T>C, p.(Leu104Pro). Segregation analysis in the parents confirmed compound heterozygosity. The variant c.1029+2T>C affects the canonical splice donor in intron 11, likely causing aberrant mRNA splicing. Skipping would result in an in-frame deletion of the 93 base pairs (bp) of the adjacent exon 11 causing a deletion of 31 AAs in the Phosphatase domain. The base pair substitution c.311T>C causes a leucine to proline missense change at the AA position 104 in the FHA domain. The splice donor variant was initially classified as likely pathogenic while the missense variant was classified as VUS (criteria: PVS1, PM2 for c.1029+2T>C, p.(?); PM2, PM3 for c.311T>C, p.(Leu104Pro)).

*This case report has been removed from the preprint version to comply with medrxiv policy. Please see the published version or contact the authors if you are interested in this information*.

Karyotype and SNP-array were normal. Panel sequencing revealed two heterozygous variants in *PNKP*, the same heterozygous splice variant c.1029+2T>C, p.(?) identified in P3 and c.151G>C, p.(Val51Leu). Sanger sequencing confirmed the presence of both variants in P4 and their heterozygosity in the parents. The canonical splice variant was inherited from one parent. The base pair substitution c.151G>C is annotated as missense change p.(Val51Leu) in the FHA domain but also affects the last nucleotide of exon 2, potentially affecting mRNA splicing. The splice donor variant was again classified as likely pathogenic while the missense variant was classified as VUS (criteria: criteria: PVS1, PM2 for c.1029+2T>C, p.(?); PM2, PM3 for c.151G>C, p.(Val51Leu)).

### Analysis of missense variants in the FHA domain

The PNKP protein consists of the N-terminal FHA domain, a Linker region, connecting the FHA domain with the central Phosphatase domain, and the C-terminal Kinase domain (4,5,25). Of 43 total unique variants reported here and in the literature (19 missense ∼ 44.2% and two in frame AA deletions ∼ 4.7%), seven missense variants (7/19 ∼ 36.8%) are located in the FHA domain, four (4/19 ∼ 21.1%) in the Phosphatase domain and eight (8/19 ∼ 42.1%) in the Kinase domain.

Mean values of the annotated CADD scores regarding the FHA (22.2), Phosphatase (23.5) and Kinase (21.5) domains are significantly higher than for the Linker domain (p < 2e-16, one-way ANOVA). These regions also contain all missense variants reported as (likely) pathogenic and disease-associated missense variants of unknown significance. The Linker region stands out with a mean CADD score of 13.8 and the lack of disease-associated missense variants.

All three missense variants identified in P1–P4 are located in the FHA domain. Review of missense variants from the literature revealed four additional disease-associated variants in this domain (Figure 2A). Beside the c.58C>T, p.(Pro20Ser) variant, which we classified as likely benign due to homozygous occurrence in the reference population (gnomAD), all missense variants previously reported as disease-associated in the FHA domain were classified as VUS using automated ACMG interpretation. Manual curation led to an evaluation as likely pathogenic for c.302C>T, p.(Pro101Leu). Three additional missense variants outside the FHA domain were also evaluated as likely pathogenic (c.526C>T, p.(Leu176Phe); c.968C>T, p.(Thr323Met); c.976G>A, p.(Glu326Lys)). Beside these four likely pathogenic missense variants, all other missense in the *PNKP* were evaluated as variants of unknown significance. Assessment based on ACMG criteria (14) of all identified disease associated *PNKP* variants can be found in File S2 (12), sheet “PNKP_variants”.

**Figure 2.**
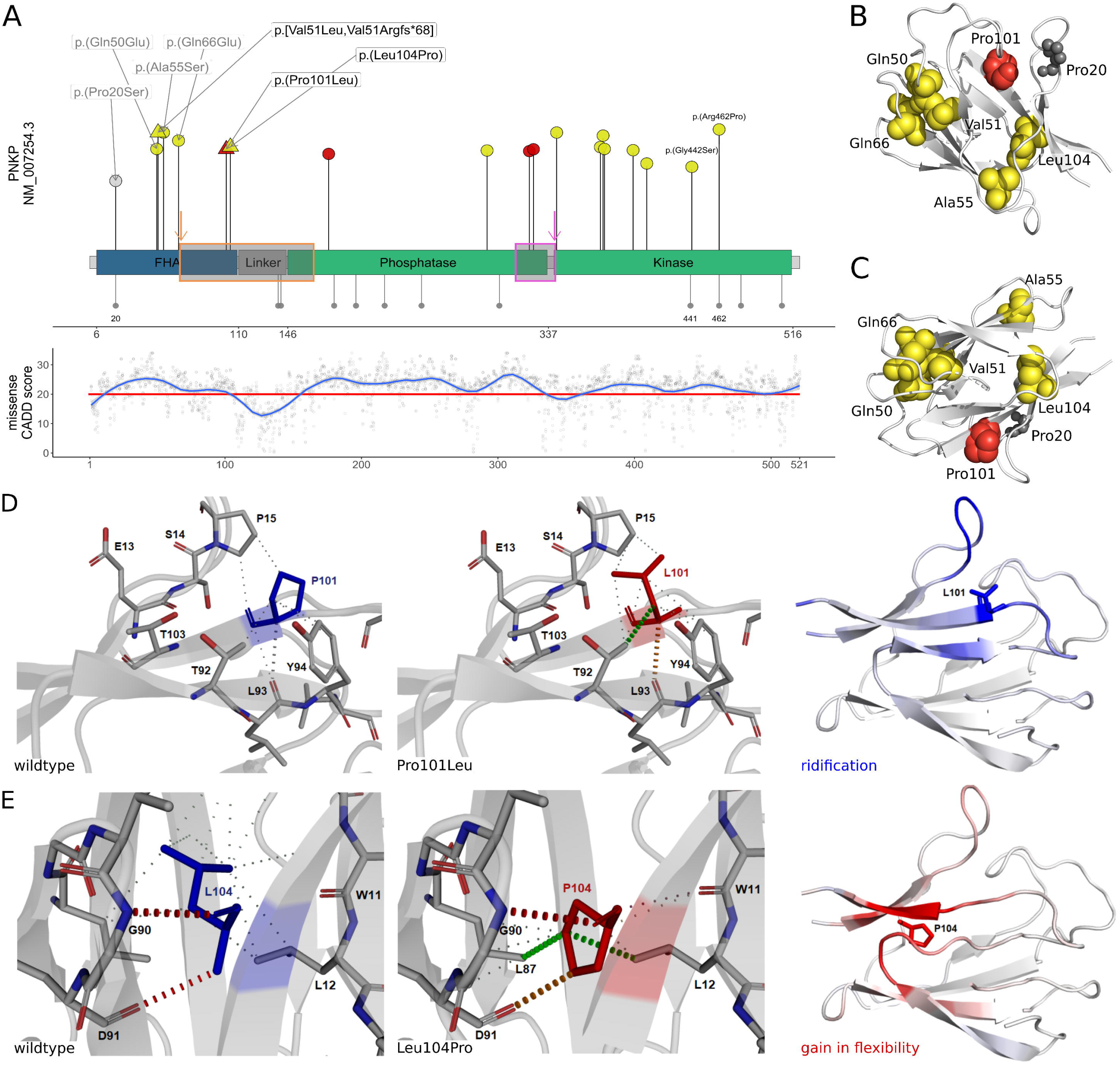
Missense variants. **(A)** Schematic depiction of the *PNKP* protein with FHA in blue, Phosphatase and Kinase domain in green and Linker region in grey (based on Uniprot Q96T60 and (20)). Disease-associated missense variants are displayed towards the top, red shapes show (likely) pathogenic variants, yellow shapes show variants of unknown significance and grey shapes show (likely) benign variants. The variants identified in the four individuals reported here are depicted as triangles (black labels), variants from the literature as dots (grey labels). The length of the segments corresponds to each variant’s CADD score. Grey dots downwards show homozygous missense variants from gnomAD, the dot size represents the logarithm of the allele count. AA positions 20, 441 and 462 are labeled for orientation. In the panel below, a generalized additive model shows the CADD values for all possible missense variants (red horizontal line = recommended cut-off (20)). Additional boxes show the affected regions of the in-frame deletions resulting from the synonymous variant c.498G>A in orange or the splicing variant c.1029+2T>C in pink. **(B)** and **(C)** Two representative views of the FHA domain crystal structure (based on 2BRF). AAs of missense variants are depicted in grey for benign variants (Pro20), in yellow for VUS (Gln50, Val51, Ala55, Gln66 and Leu104) and in red for pathogenic variants (Pro101). All missense variants classified as unknown significance or (likely) pathogenic affect conserved residues in beta-sheets. **(D)** The substitution Pro101Leu alters the local interatomic interaction of the protein, generating important steric clashes at the same time it decreases molecule stability. The most right panel represents the ΔVibrational Entropy Energy derived from the Pro101Leu variant, showing a local rigidification of the protein. **(E)** The Leu104Pro variant generates the opposite effect observed for Pro101Leu as it introduces a proline in the core of the FHA domain, destabilizing the protein domain. The most right panel shows the ΔVibrational Entropy Energy with a local gain in flexibility.

**Figure 3.**
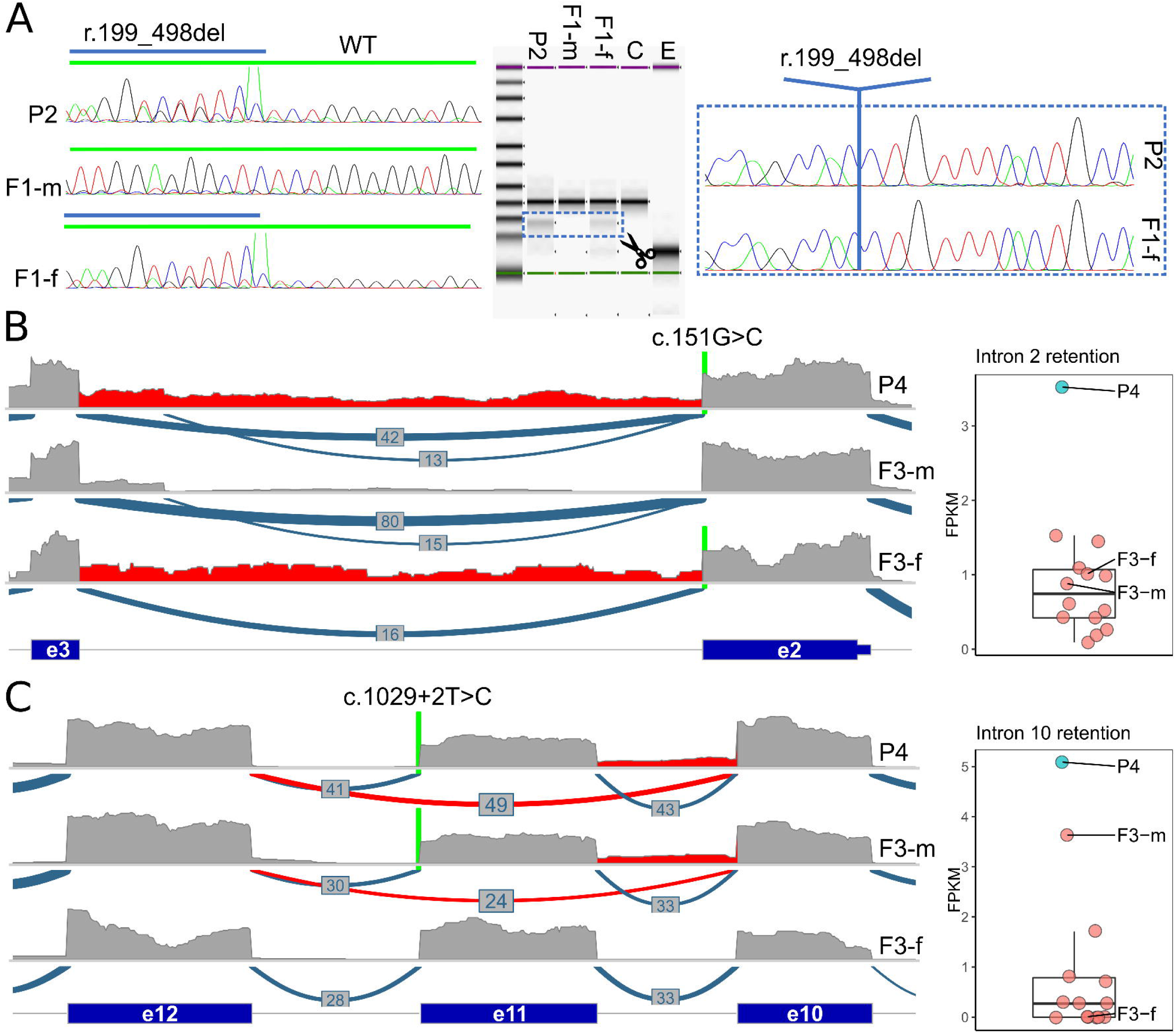
RNA analyses. **(A)** RT-PCR and subsequent Sanger-sequencing (left panel) on cDNA derived from RNA of individual P2 and both parents (F1-m, F1-f) to determine the effect of the c.498G>A variant. In P2 and the heterozygous carrier parent a smaller product is present (middle panel). (right panel) Skipping of exon 4 (r.199_498del) causes an in-frame deletion on protein level (p.Leu67_Lys166del). WT: wildtype, C: control, E: empty. **(B)** Coverage and Sashimi-plot (left panel) of RNA-seq from family 3 for the exon 2 – exon 3 region. The c.151G>C variant is indicated with a green line. Coverage plot for P4 and the heterozygous carrier parent (F3-f) (highlighted in red), indicating intron 2 retention (r.151_152ins151+1_152-1). This is confirmed by the FPKM values shown as box- and scatter plots (right side), where P4 is the only outlier. **(C)** Coverage and Sashimi-plot (left panel) of RNA-seq data from family 3 for the exon 10 – exon 12 region. The position of the c.1029+2T>C variant is indicated with a green line. The Sashimi-plot shows skipping of exon 11 (r.937_1029del; highlighted in red) in P4 and the heterozygous carrier parent (F3-m), which is predicted to cause an in-frame deletion (p.Phe313_Pro343del). Additionally, the coverage plot indicates retention of intron 10 (r.936_937ins936+1_937-1; red highlight) in P4 and F3-m, which is confirmed by the FPKM values for this exon (right panel and predicted to cause a frameshift (p.Leu313Valfs*16).

The spatial distribution of the AA residues affected by missense variants in the crystal structure of the FHA domain (based on 2BRF from RCSB Protein Data Bank (PDB) showed that most disease-associated missense variants affect conserved residues in beta-sheets (Figure S3A). Analysis with mutation3D revealed clustering of the affected AA positions 50, 51, 55, 66, 101 and 104 in the FHA domain with a significant p-value (bootstraping) of 0.0112 (Figure 2B and 2C).

According to Missense3D, the c.302C>T, p.(Pro101Leu) variant triggers a local steric clash alert (Figure 2D). Additionally, based on the DynaMut web server predictor, the effect of this variant is stabilizing and the ΔVibrational Entropy Energy between wildtype and mutant structure is predicted to slightly decrease molecule flexibility (Figure 2D). The c.311T>C, p.(Leu104Pro) substitution introduces a buried proline in the core of the protein domain, which tends to be particularly damaging with its restricted backbone conformation (Figure 2E). In fact, according to Dynamut, the variant has the opposite effect of p.(Pro101Leu) and is predicted to be destabilizing with increase of the molecule flexibility (Figure 2E). Comprehensive results of computational analyses for all variants in the FHA domain are listed in Table 2.

**Table 1.** Identified *PNKP* variants.

|  | P1& P2 | P1&P2 | P3 | P3&P4 | P4 |
| --- | --- | --- | --- | --- | --- |
| **chromosomal position** | chr19:50368580G>A | chr19:50368384C>T | chr19:50368571A>G | chr19:50365626A>G | chr19:50370311C>G |
| **c-code\*** | c.302C>T | c.498G>A | c.311T>C | c.1029+2T>C | c.151G>C |
| **r-code** |  | r.199_498del |  | r.[937_1029del,936_937ins936+1_937-1] | r.[151g>c,151_152ins151+1_152-1] |
| **p-code\*\*** | p.(Pro101Leu) | p.Leu67_Lys166del | p.(Leu104Pro) | p.[Phe313_Pro343del,Leu312_Phe313ins*18] | p.[Val51Leu,Val51Argfs*68] |
| **effect** | missense variant | splice region variant& synonymous variant | missense variant | splice donor variant& intron variant | missense variant& splice region variant |
| **InterVar** | Uncertain significance | Uncertain significance | Uncertain significance | Pathogenic | Uncertain significance |
| **ACMG** | Likely pathogenic | Likely pathogenic | Uncertain significance | Likely pathogenic | Uncertain significance |
| **evidence** | PS4_MOD;PM1_SUP;PM2_SUP;PM3;PP3 | PVS1_STR;PM1_SUP;PM2_SUP | PM2_SUP;PM3;PP3 | PVS1_STR;PM3_SUP;PS4_MOD;PM2_SUP | PM2_SUP;PM3;PP3 |

\*reference transcript: NM\_007254.3
\*\*p-codes without round brackets are after RNA analyses

**Table 2.** Structural modelling results for missense variants in the FHA domain.

| p-code* | p.(Gln50Glu) | p.[Val51Leu,<br>Val51Argfs*68] | p.(Ala55Ser) | p.(Gln66Glu) | p.(Pro101Leu) | p.(Leu104Pro) |
| --- | --- | --- | --- | --- | --- | --- |
| case or reference | PMID: 31167812 | P4 | PMID: 24965255 | PMID: 30956058 | P1&P2 | P3 |
| CADD<br>PHRED<br>score v1.6 | 26.5 | 22.7 | 25.0 | 26.0 | 27.9 | 28.5 |
| Missense3<br>D | replaces a buried uncharged residue with a charged residue | no structural damage | no structural damage | replaces a buried uncharged residue with a charged residue | local steric clash alert | introduces a buried proline in the core of the protein domain |
| DynaMut | destabilizing ($\Delta\Delta G$: -0.362 kcal/mol)<br>increase of molecule flexibility ($\Delta\Delta SVib$ ENCoM: 0.357 kcal/mol*K) | destabilizing ($\Delta\Delta G$: -0.670 kcal/mol)<br>little decrease of molecule flexibility ($\Delta\Delta SVib$ ENCoM: -0.591 kcal/mol*K) | stabilizing ($\Delta\Delta G$: 0.989 kcal/mol)<br>little decrease of molecule flexibility ($\Delta\Delta SVib$ ENCoM: -0.345 kcal/mol*K). | stabilizing ($\Delta\Delta G$: 0.294 kcal/mol)<br>very little increase of molecule flexibility ($\Delta\Delta SVib$ ENCoM: 0.084 kcal/mol*K) | stabilizing ($\Delta\Delta G$: 1.510 kcal/mol)<br>slightly decrease of molecule flexibility ($\Delta\Delta SVib$ ENCoM: -0.287 kcal/mol*K) | destabilizing ($\Delta\Delta G$: -1.608 kcal/mol)<br>increase of molecule flexibility ($\Delta\Delta SVib$ ENCoM: 0.615 kcal/mol*K) |

\*reference transcript: NM\_007254.3

### RNA analyses

To determine the effect of the parentally inherited c.498G>A variant in P1 and P2, we performed RT-PCR and Sanger-sequencing on cDNA from RNA of individual P2 and both parents (F1-m, F1-f). In addition to the wildtype product present in all three samples, we identified a second, smaller PCR product in the sample of one parent and P2. Sanger-sequencing of this smaller product revealed skipping of the 30 base pair long exon 4 (r.199_498del) leading to an in-frame deletion of the 100 AAs of exon 4 on protein level (p.Leu67_Lys166del) (Figure 3A).

To elucidate the effect of the base pair substitution c.151G>C, affecting the last base of exon 2, and proof aberrant splicing of the canonical donor variant c.1029+2T>C, we performed RNA sequencing from PAXgene blood sample of P4 and both parents (Figure 3). RNA sequencing data in the sample of P4 and the heterozygous carrier parent (F3-f) showed mainly normal splicing events for the exon 2/3 region with nucleotide exchange c.151G>C leading to the missense change p.Val51Leu one consequence from this allele. The sequencing reads also support an aberrant transcript with retention of intron 2 (r.151_152ins151+1_152-1), which is predicted to cause a frameshift and a premature stop codon (p.Val51Argfs*68). Overall the c.151G>C base exchange causes a complex effect on RNA and protein level (p.[Val51Leu,Val51Argfs*68]) (Figure 3B). Estimating the predominant aberrant transcript and effect is complicated by the presence of the second variant in the index and apparently incomplete nonsense mediated RNA-decay. Based on the allele fraction of the RNA-seq reads with the c.151G>C change (10/35 overall), the two consequences from this allele (4 correctly spliced with base exchange, 6 with retention) are about equally expressed.

For the variant c.1029+2T>C inherited from one parent (F3-m) our RNA-seq data confirmed the expected effect of exon 11 skipping (r.937_1029del) leading to an in-frame deletion of the 31 AAs (p.Phe313_Pro343del). We identified evidence for an intron 10 retention (r.936_937ins936+1_937-1) as a second aberrant transcript, which is predicted to cause a frameshift (p.Leu313Valfs*16) (Figure 3C). Thus, also this canonical splice variant causes a complex mixture of aberrant transcript (p.[Phe313_Pro343del,Leu313Valfs*16]). Estimating the predominant effect is again complicated by multiple novel transcripts from both alleles in the index. From the spliced reads observed over the skipped exon in one parent (compare Figure 3C) one would calculate an exon-inclusion ratio (or percent spliced in; PSI) of 56.8% (((30+33)/2)/((30+33)/2 + 24)) for exon 11. To restore a model with equal expression of the transcripts from both alleles and assuming no nonsense mediated decay, 7.5 transcripts would have to be assigned to the retention event (add 7.5 to the denominator) which would constitute ∼11.9% (7.5/63) of all transcripts. This calculation is in agreement with the observed coverage profile between exon 10 and intron 10 (11/69 ∼15.9%). Overall, this confirms that the predominant aberrant splice effect caused by the c.1029+2T>C variant is in-frame exon skipping.

## DISCUSSION

Before our report, *PNKP* was already associated with a remarkably wide phenotypic disease spectrum. This could be related to the protein’s multi domain architecture. The FHA domain recruits the protein to sites with DNA damage (26), where it is involved in repair of both single- and double-strand breaks through its dual enzymatically active Kinase and Phosphatase domains (25,27).

Despite the various *PNKP*-associated phenotypes (5–9,38–41) prenatal presentations in humans, especially noticeable concomitant brain anomalies, were unreported to date. Our compilation of prenatal diagnostic procedures and fetal pathological examination of P1 and P2 revealed neurodevelopmental and neurodegenerative brain alterations comparable to those described in mouse models with neuronal tissue-specific inactivation of *PNKP* (32). These include general hypoplasia of different cerebral and cerebellar regions, without a histologically recognizable neuronal migration disorder. Furthermore, the two fetuses of family 1 show a convincing phenotypic accordance concerning enlarged ventricles, thin corpus callosum and microcephaly with the previously described MCSZ phenotype during infancy (6). While the exact pathomechanism causing microcephaly in some *PNKP-* associated phenotypes is still disputed, the prenatal manifestation represents the most severe clinical outcome. This may arise from extreme genome instability in neurons with impaired development itself on the one hand and increased cell death on the other hand, the latter one may lead to the extensive white matter deficit reported for P1.

With suspicion of recurrent microcephaly, fetal autopsy of P2 was oriented towards an underlying syndromic disorder. Missing macroscopic signs for holoprosencephaly such as hypotelorism, hypoplastic anterior cranial fossae or absent cribriform plate of ethmoid bone in the first pathological examination (limited on microscopic level due to autolysis) together with the unremarkable holoprosencephaly panel analysis provide sufficient arguments to retrospectively rule out holoprosencephaly in P1. The synergy of unbiased exome sequencing for prenatal anomalies and exact phenotyping in a syndrome-oriented fetal autopsy in P2 highlight their importance for diagnosing an unusual manifestation of a known disease and evaluation of a novel variant of uncertain clinical significance.

Given the OFC values observed immediately after birth in P3 and P4, the presence of prenatal microcephaly in these individuals is obvious. The diagnosis of PM relies on ultrasound (US) measurements in comparison to distributions for the respective gestational age and exclusion of exogenous causes such as infectious diseases. Until the recent Zika virus outbreak, no international standards and guidelines had been defined and research on the diagnostic performance of US measurements for fetal microcephaly was hampered by the overall rarity of the condition (33). Performance for prenatal US diagnosis seems good at the more extreme ends (< -4 SD) or when additional brain anomalies are present (33), like in the case of the two fetuses of family 1. Additionally, improved US technology and specialist training during the last years might have enabled these prenatal diagnoses.

We recommend interpreting the phenotypic presentations associated with biallelic variants in *PNKP* as continuous spectrum instead of the separated clinical entities MCSZ, AOA4 and CMT2B2, in agreement with previous suggestions (28). In this disease model, our report adds the most severe, prenatal presentation with extreme microcephaly and possibly unviable additional cerebral anomalies in the two sibling fetuses. With regard to the grade of severity, this is followed by postnatal microcephaly with severe to mild intellectual disability combined with further neurological symptoms like epilepsy and finally by adult-onset polyneuropathy as the mildest presentation (Figure 4). Observing this clinical variability, the question arises whether there could be a relation to genetic variant context. On a genomic level, *PNKP* stands out given the specific composition with many small introns towards the 3’ end when compared to the average human intron size (34). Smaller introns are associated with a higher likelihood of intron retention (35), a mechanism we proved for two novel variants here. RNA analyses of disease-associated *PNKP* variants were previously not reported, despite many intronic and splice sites affecting changes described (6,26,28,29,36–43). The results of our RNA splicing analyses for one novel silent variant, one known splice donor variant and moreover one variant annotated as missense point toward a likely underappreciated pathomechanism. Predictions by computational tools implicate a splice effect for 15 of 43 (35%) *PNKP* variants, which implies a need for further functional evaluation. We show that even complex splice events in *PNKP* can readily be assessed with RT-PCR or RNA-seq analyses from peripheral blood. Expanding these analyses to newly identified variants will support variant pathogenicity interpretation and improve understanding of aberrant splicing for genes with similar exon configuration.

**Figure 4.**
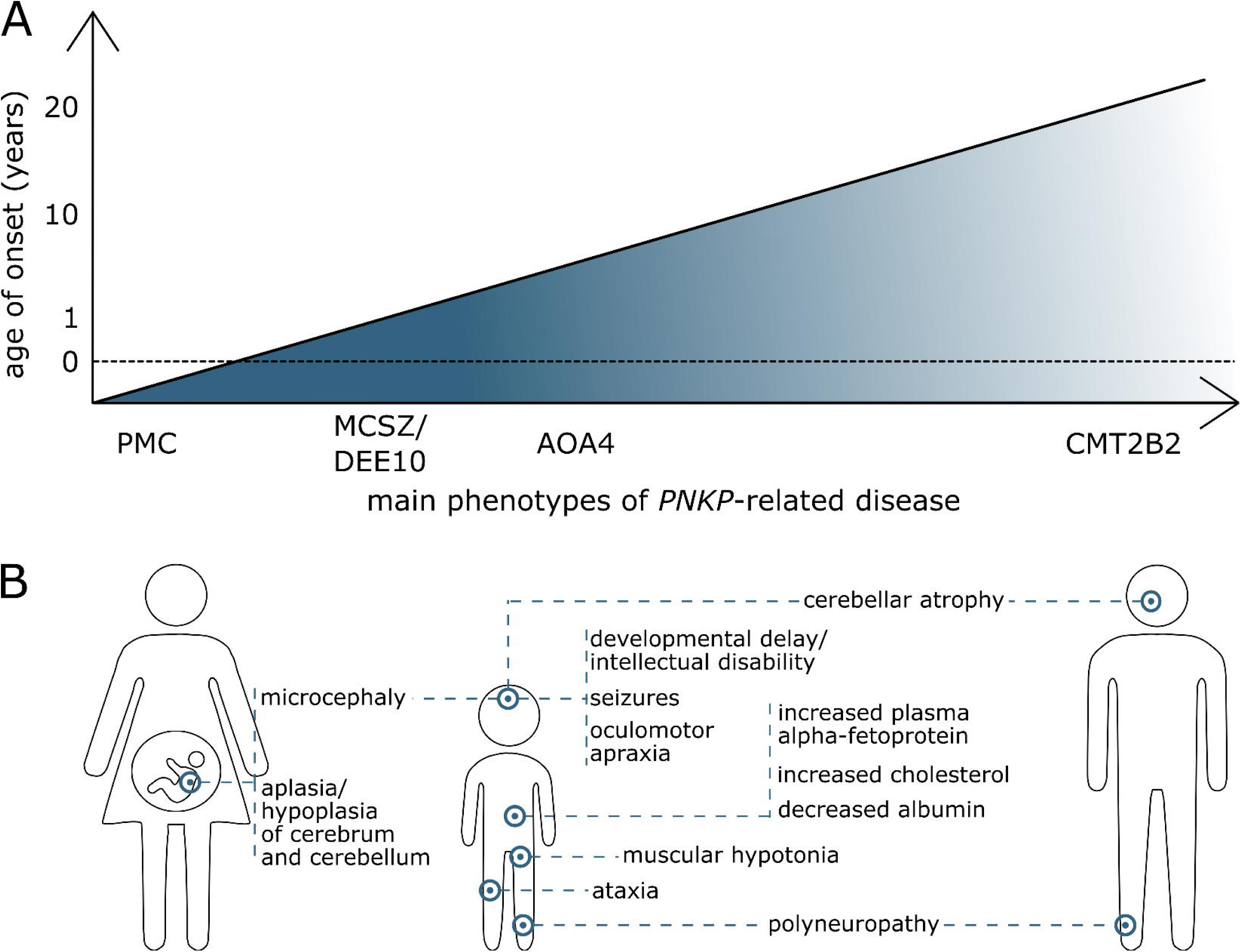
Phenotypes. **(A)** Schematic indicating the proposed phenotypic continuum in *PNKP*-related disease: MCSZ (microcephaly, seizure and developmental delay; MIM# 613402) and developmental and epileptic encephalopathy (DEE10; MIM# 613402), AOA4 (ataxia-oculomotor apraxia type 4; MIM# 616267) and Charcot-Marie-Tooth disease, type 2B2 (CMT2B2; MIM# 605589). Prenatal manifestation is added as prenatal microcephaly (PMC). Interpreted severity of the disease is shown by color course. **(B)** Comparison of the main clinical aspects between prenatal period (left), childhood (middle) and adulthood (right). Aplasia/hypoplasia of the cerebrum (HP:0007364) and the cerebellum (HP:0007360) is exclusively described in the prenatal period. Microcephaly (HP:0000252) is one of the main features of the prenatal and childhood phenotype MCSZ. MCSZ and DEE10 also comprise developmental delay/intellectual disability (HP:0001263; HP:0001249), seizures (HP:0001250) and muscular hypotonia (HP:0001252). Ataxia (HP:0001251) and oculomotor apraxia (HP:0000657) are characteristic for AOA4. Distinct biochemical features (elevated alpha-fetoprotein HP:0006254, hypoalbuminemia HP:0003073, hypercholesterolemia HP:0003124) were mostly described in cases with AOA4. Overlap between childhood and adulthood presentation are cerebellar atrophy (HP:0001272) and polyneuropathy (HP:0000763).

Next to the often complex and possibly hidden splice effects identified here, true AA substitutions are to date the most difficult variants to interpret. Evaluation of missense variants can be composed of evolutionary conservation, functional predictions and genetic context as it is used comprehensively e.g. in the CADD score (44). Here we complemented the analysis of such scores in the linear protein model with structural protein modelling. Four variants identified in the three families affect the FHA domain. The AA residues affected by the missense variants p.(Gln50Glu), p.[Val51Leu,Val51Argfs*68], p.(Ala55Ser), p.(Gln66Glu), p.(Pro101Leu), and p.(Leu104Pro) show significant clustering with other AA changes described in the literature. Interestingly, the compound heterozygous variants identified in family 1 alter the same region of PNKP: the variant p.Leu67_Lys166del predominantly results in a deletion affecting parts of the FHA, Linker and Phosphatase domain and thereby encompasses a total of around 20% of PNKP, while the second is the missense variant p.(Pro101Leu) located at the C-terminus of the FHA domain. Previously, no disease-associated variants were described in this particular PNKP region, which shows high conservation and absence of homozygous variants in population databases. The severity of the observed prenatal phenotype may either be generated by the combined interdomain effect of the large in frame deletion or serves as an intriguing confirmation of a “Wald’s domain” at the crossing between the FHA and Linker domains, which has recently been proposed as a survivorship bias for the disease associated variants reported in *PNKP* (11).

Taking into account the different mutational mechanisms of *PNKP* variants and the wide phenotypic spectrum, currently no clear genotype-phenotype correlation and therefore no accurate phenotypic prediction in association to a specific variant seems possible. While *PNKP* may be an extreme example, this is a common challenge in disorders with autosomal-recessive inheritance. In homozygous state, either because of consanguinity or due to founder variants, other effects of the haplotype can influence phenotype, while in compound heterozygous state the combination of two variants with possibly different effects each complicates accurate phenotype association. The latter is exemplified by the difference in clinical severity in individuals P3 and P4 who both carry the same splice variant c.1029+2T>C causing a mixture of protein effects (p.[Phe313_Pro343del, Leu312_Phe313ins*18]) on one allele and a different missense variant each on the second allele. Individual P4, who has the milder phenotype without seizures, carriers the splice region variant c.151G>C causing mixture of missense (p.Val51Leu) and truncating effect (p.Val51Argfs*68). In contrast, individual P3 who has severe neurodevelopmental delay, seizures, and extreme microcephaly (< -6 SD) carries the missense variant c.311T>C, p.(Leu104Pro), which lies in the same FHA region and close proximity of the variants in family 1. While this anecdotal correlation seems convincing, dysfunction of the phosphatase domain was already associated with *PNKP*-associated neurodevelopmental phenotype (11,26), which we can support with variants in all three families affecting this domain. Systematic multi-level functional studies matched with standardized (HPO; see Figure 4) clinical assessment will be needed to reach precise genotype-phenotype prediction.

Here, we extended the “*PNKP*-associated disorder continuum” to the prenatal period and complemented missense variant interpretation with 3D structure analysis and presented the first RNA-analyses known so far for *PNKP* variants. Future studies will need to combine these techniques with detailed phenotyping taking into account the *PNKP*-continuum and ideally add massive parallel functional tests. Due to the associated genetic heterogeneity the diagnosis of primary microcephaly requires (trio-) exome sequencing. The knowledge of distinct fetal phenotypes will be helpful for genetic variant assessment, especially those with unknown significance. Only with knowledge of variant pathogenicity and expected symptoms, we will be able to improve counselling in the prenatal setting, management in the postnatal period, prenatal diagnosis in subsequent pregnancies, and finally enable potential evaluation of treatment in the future.

## Data Availability

All data generated or analyzed during this study can be found in the online version of this article at the publisher's website with access to supplementary data resources or on Zenodo.

## ACKNOWLEDGEMENTS

We thank all involved families for participating in this study. We thank Viktoria Wischmann (medical-laboratory assistant, Institute of Pathology, Philipps University of Marburg) for engaged collaboration and excellent technical support in performing RNA extraction for P2 as well as Mirko Wegscheider (Department of Neurology, University of Leipzig) for his kind beneficial assistance with interpretation of MRI images.

## AUTHORS’ CONTRIBUTIONS

B.P. conceived the initial study concept, S.N. and I.K. coordinated collection of clinical and genetic data through matchmaking and personal communications. S.N. and I.K. reviewed literature data and standardized the clinical HPO terms. I.K., A.S., R.A.J., T.B., J.D., S.S., M.P., S.R., C.R., H.R. and K.S. provided clinical and genetic data and performed clinical assessments. K.S. and H.R. performed fetal pathology analysis. K.S. provided fetal pathological images and fetal RNA samples. S.Sc. conducted RT-PCR analyses. D.L.D., M.R. and B.P. analyzed RNA-seq data. L.B.G., S.N., B.P. and A.L. performed structural protein analysis. S.N., A.L., L.B.G. and B.P. created main figures and the Supplementary materials. S.N., I.K. and B.P. wrote and edited the manuscript. All authors reviewed, commented and agreed on the final draft manuscript.

## FUNDING

B.P. is supported by the Deutsche Forschungsgemeinschaft (DFG) through grant PO2366/2–1. D.L.D. is funded through “Clinician Scientist Programm, Medizinische Fakultät der Universität Leipzig”.

## COMPETING INTEREST

All authors involved in the study declare no conflicts of interest relevant to this study.

## WEB RESOURCES

gnomAD browser: http://gnomad.broadinstitute.org/

Mutalyzer: https://mutalyzer.nl

ClinVar: https://www.ncbi.nlm.nih.gov/clinvar/

Missense3D: http://missense3d.bc.ic.ac.uk/missense3d/

Dynamut: http://biosig.unimelb.edu.au/dynamut/

Mutation3D: http://mutation3d.org/

## ABBREVIATIONS

AA: amino acid
ACMG: American College of Medical Genetics and Genomics
GA: gestational age
HPO: Human Phenotype Ontology
MRI: magnetic resonance imaging
PNKP: Polynucleotide kinase 3’-phosphatase
SD: standard deviation

## Notes

### Competing Interest Statement

The authors have declared no competing interest.

### Author Declarations

The project was approved by the ethic committee of the University of Leipzig, Germany (224/16-ek and 402/16-ek) and was conducted in concordance to the declaration of Helsinki.

