## Supplementary notes for "Prenatal phenotype of *PNKP*-related primary microcephaly associated with variants in the FHA and Phosphatase domain"

### SUPPLEMENTARY METHODS

#### Genetic Analyses and Variant Annotation

Genomic DNA was extracted using standard methods either from peripheral blood samples of P3, P4, and all concerned parents or from an amniotic fluid sample for P1 and P2. Enrichment for exome sequencing of P2 and both his parents was done using the Twist Human Core Exome Probes (Twist Bioscience, San Francisco, USA) and resulting libraries were sequenced on an Illumina NextSeq 500/550 platform (Illumina Inc., San Diego, USA). Between 99.0 % (index) and 98.7 % of the targeted sequences reached at least 20-fold coverage. Clinical exome sequencing of P3 and P4 (TruSight One Panel v1, Illumina Inc., 4813 genes) was performed on an Illumina NextSeq 500/550 platform (Illumina, Inc., San Diego, USA); 97.8% (P3) and 97.3% (P4), respectively, of the targeted sequences reached at least 20-fold coverage. The resulting raw sequencing data was processed using the cloud based “Varfeed” pipeline and resulting variants were analyzed with the browser based “Varvis” genomics software (Limbus Medical Technologies GmbH, Rostock, Germany).

Compound heterozygosity and segregation was confirmed by Sanger sequencing. The following primer pairs were used: exon 2 (P3) 5’- AGCAGTTAATGGTGGGGAAA-3’ and 5’- AAGCGTCCCTCTGGATTGTT-3’; exon 4 (P1, P2, P4) 5’- CCAAAGCCCGTTCCAAAGTG-3’ and 5’- TTGAGAGCACGCAACAAACG-3’; exon 11 (P3, P4) 5’- GCCTGTGTCTGATGTTCGTC-3’ and 5’-GAATCCCACTGCGACAACC-3’.

VariantValidator (version 1.0.4.dev120+g683823d) was used to convert variants into VCF format (1) and consecutively annotated with scores from the dbNSFP database (version 2.93) (2) using SnpEff/SnpSift (v4.3.1t) (3,4). CADD score values (version 1.6) (5) were annotated through the online platform using the VCF as input. Splicing prediction was done via SPIDEX/SPANR (SPIDEX/SPANR: http://tools.genes.toronto.edu) (6) and dbscSNV (http://www.liulab.science/dbscsnv.html) scores (2) and annotated using SnpSift.

#### Detailed fetal autopsy

Fetal pathological examination of P1 and P2 included anthropometric measurement, careful inspection of the external characteristics, in particular of the facial aspect, X-ray examination (fetogram) before opening of the body cavity, removing of the organs, magnifier-assisted macroscopic preparation of the organs (LUXO WAVE PLUS magnifier lamp 3.5 dpt, Glamox Luxo Lighting GmbH, Hildesheim, Germany), microscopic examination and separate neuropathological examination of the central nervous system. All steps of examination resp. autopsy were documented photographically using a repro stand with a motorized heightadjustable camera mount (Kaiser Zootechnics GmbH & Co. KG, Buchen, Germany). All photographs were taken with a digital camera (Canon Power Shot G5; Canon, Tokyo, Japan) and images were further processed for exposure, color saturation and image size (Adobe Photoshop; Mountain View, CA, USA).

**RT-PCR in family 1**

The PAXgene Blood System (Becton Dickinson, Franklin Lakes, NJ) was used to extract RNA from peripheral blood lymphocytes of both parents from family 1. RNA was processed with DNase I (Qiagen, Hilden, Germany) and transcribed into cDNA with the Superscript II Reverse Transcriptase Kit (Invitrogen, Carlsbad, CA) according to manufacturer's protocol. RT-PCR on cDNA from family 1 was performed with three different primer pairs, spanning exon-exon junctions and the c.498G>A PNKP variant (P1f: 5′-ACTCAAGTGGAGCTGGTCGC-3′ and P1r: 5′-GAAAGACCTTCCCAGAGCGT-3′ (amplicon length: 412bp); P2f: 5′-AGATCCTGAGACCCGGACAG-3′ and P2r: 5′ GGGTACAAGATCCTCCAGTCAC-3′ (amplicon length: 426bp)). Resulting PCR products were visualized by gel electrophoresis and subsequently Sanger-sequenced.

**RNA-seq in family 3**

Libraries were prepared from PAXgene RNA (PAXgene Blood RNA Kit v2; Qiagen, Hilden, Germany) using the TruSeq RNA Library Prep Kit v2 and 500ng total RNA according to the manufacturer's instructions. Libraries were assessed for quality using the Agilent 2200 TapeStation System and for quantity using the Qubit Fluorometric quantification (Thermo Fisher Scientific Inc., Waltham, USA). All libraries presented a fragment length distribution with peaks at about 280bp. Sequencing was performed on an Illumina NextSeq platform using 76bp paired-end reads. After demultiplexing, adapters were trimmed using leehom (7). We performed a quality check using FastQC (https://www.bioinformatics.babraham.ac.uk/projects/fastqc/). Cutadapt (8) was used to remove overrepresented reads. These such reprocessed reads were mapped to the hg38 reference using STAR aligner (9) version 2.6.1.d. On average 126.92 ± 37.43 million reads were mapped per library. Alignment files were sorted and indexed with samtools (10). Final BAM files were visualized as trio and with controls from the same run and inspected for alternative splicing using the IGV browser (11) and its inbuilt Sashimi-plot functionality as described previously (12). As the visual analysis indicated intron retention events, we applied the recently published iREAD command line tool (13) version 0.8.5 and a BED region file for

PNKP introns (reference transcript ENST00000322344.8; generated using UCSC Galaxy (https://usegalaxy.org/)) to quantify the PNKP intron retention events in all RNA-seq samples from the run. Resulting FPKM (Fragments per Kilobase of transcript per Million) values were loaded into R (version 4.0.3), filtered (packages "tidyverse" and "readxl") and visualized as box and scatter plots using the "ggplot2" package. Additionally, BAM files were loaded into R to visualize spliced alignments and retention events using the "Gviz" and "trackViewer" libraries. Inkscape 1.0.1 was used to adjust Figure 3 for parts which could not be directly composed in R.

**Analysis of linear missense variant distribution**

Disease-associated variants were plotted onto the linear protein structure of PNKP using R (version 4.1.0) with the package ggplot2. Domain information from UniProt (14) was complemented with the Linker domain from literature (15). Variant distribution was visually investigated and compared to locations of missense variants in *PNKP*, which are found in the gnomAD database in homozygous state (16). We then analyzed protein regions constrained for missense variation by generating all possible *PNKP* missense variants, annotating them with the computational score CADD PHRED v1.6 (5). The results were plotted by amino-acid position and fitting a generalized additive model ("geom_smooth" function in ggplot2). CADD PHRED score values of all annotated domains were compared via one-way ANOVA test using R ("aov" function).

**3D clustering analysis and structural modelling**

Disease-associated missense variants at amino acid (AA) positions 50, 51, 55, 66, 101 and 104 of the FHA domain were highlighted using the crystal structure 2BRF (17) downloaded from RCSB Protein Data Bank (PDB). Variant clustering was analyzed with the command line version of mutation3D (18) (settings: complete linkage-distance 20, protein length 110, number of bootstrapping iterations 10,000) using 2BRF from PDB as input model. Additionally, the Missense3D (19) and the Dynamut server were used to analyze the impact of missense variants on protein dynamics and stability. Final molecular graphics of tertiary structures were visualized and rendered using the PyMOL software (Version 2.4.0; Schrödinger LLC, New York, USA).

**Multiple Sequence Alignment and interaction with XRCC1-derived phosphopeptide**

PNKP protein sequences were obtained from UniProt (14). Multiple Sequence Alignment (MSA) was performed with the EMBL-EBI's server for Multiple Alignment using Fast Fourier Transform (MAFFT) (20) as it is recommended for better accuracy (21). The MSA was visualized and analyzed in Jalview 2.11.1.2 (https://www.jalview.org/). Jalview determined the conservation score based on the alignment. The crystal structure of the FHA Domain of PNKP was obtained from the Protein Data Bank (PDB ID: 2BRF/2W3O) (17).

**Additional data**

We provide comprehensive clinical data and raw data on variant lists as online dataset on Zenodo (22).

### SUPPLEMENTARY FIGURES

#### Figure S1 | Postnatal percentile curves of head circumference

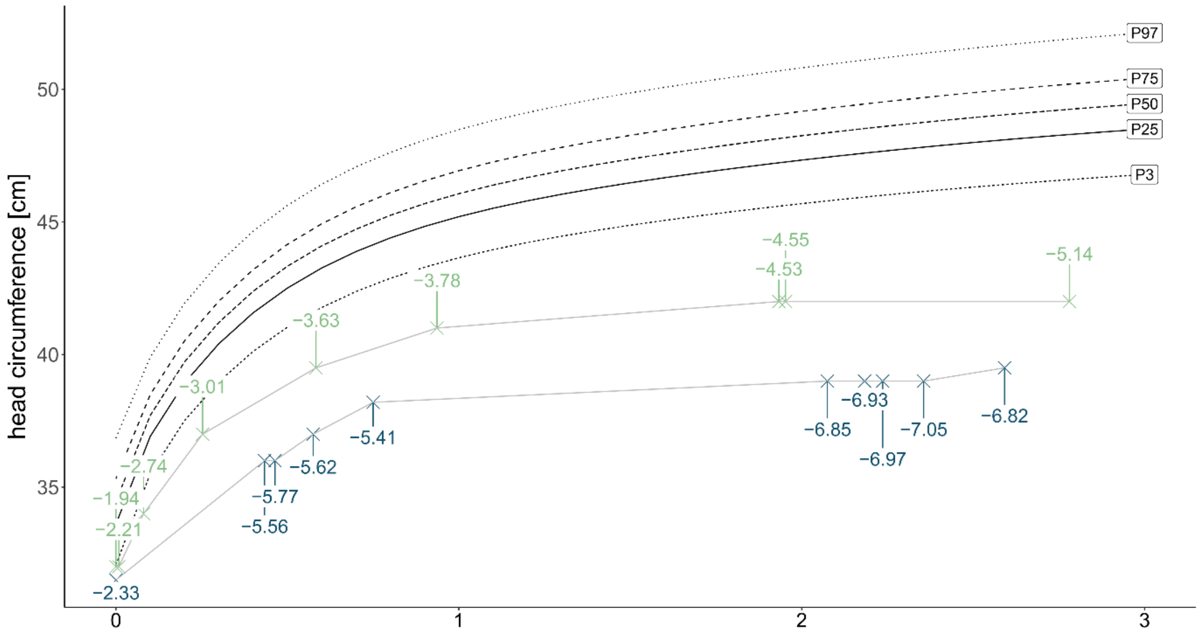


Postnatal percentiles of the head circumference for P3 (blue) and P4 (green) compared to WHO percentiles for P3, P25, P50, P75 and P97. Both curves show progressive primary microcephaly.

**Figure S2 | Postnatal cMRI**

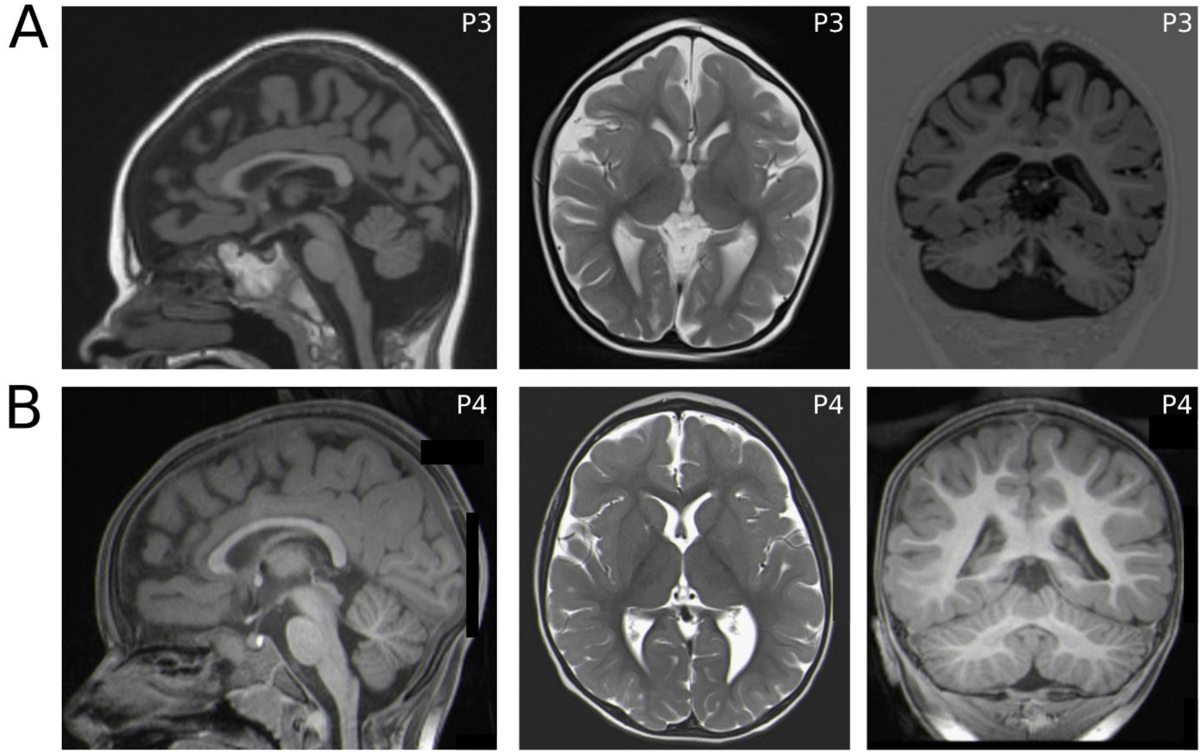


**(A)** cMRI sagittal T1 (left panel), transversal T2 (middle panel) and frontal T1 (right panel) of individual P3 years showed a microcephaly with simplified gyral pattern, small corpus callosum, supra- and infratentorial deficit of white matter and substance deficit of the cerebellum. **(B)** cMRI sagittal T1 (left panel), transversal T2 (middle panel) and frontal T1 (right panel) of individual P4 years showed microcephaly without additional brain abnormalities.

**Figure S3 | Multiple Sequence Alignment and structural analysis of interaction with XRCC1-derived phosphopeptide**

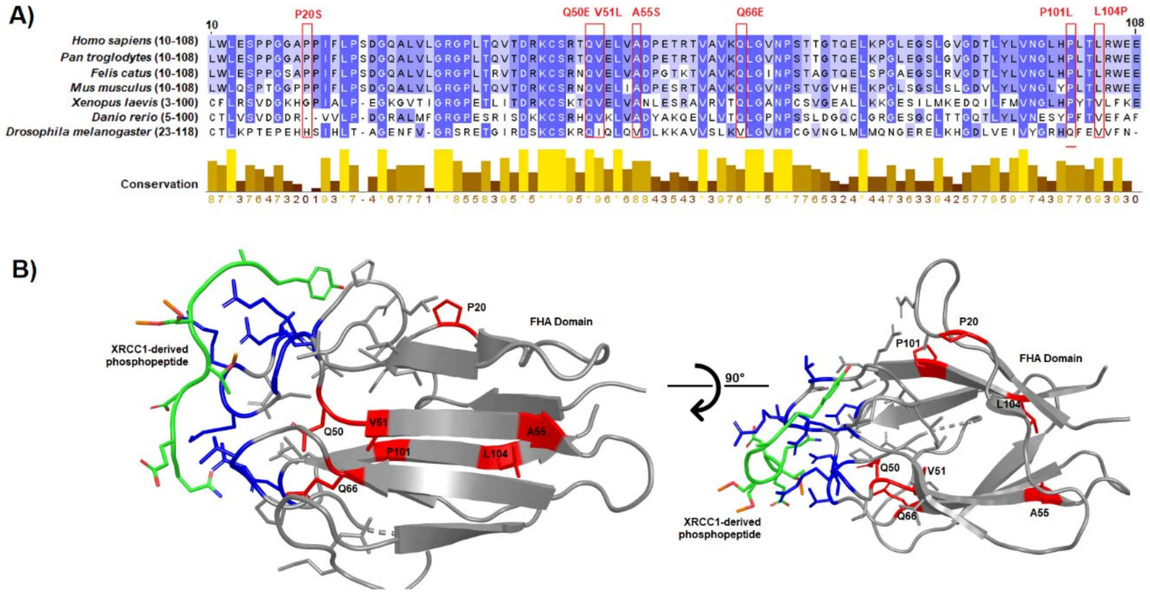


**(A)** MSA of the FHA domain between several species. Amino acids are colored according to their percentage identity. Variants reported in the FHA domain are highlighted in red. **(B)** Structural location of missense variants (red) in the FHA domain of PNKP (PDB ID: 2W3O). The amino acids interacting with the XRCC1-derived phosphopeptide (in green) are colored in blue. None of the variants appear to affect the protein-protein interaction interface.
